# Only Anxiety Remains Reliably Associated with Paediatric Mild Traumatic Brain Injury at Two Years Follow-up After Adjusting for Pre-Existing Mental Health

**DOI:** 10.1101/2024.03.22.24304723

**Authors:** Grace Revill, Norman Poole, Christina Carlisi, Anthony S. David, Vaughan Bell

**Author notes:** Corresponding Author: Prof Vaughan Bell, 1-19 Torrington Place, London, WC1E 7HB.

## Abstract

**Background:** Evidence that mild traumatic brain injury (mTBI) causes psychiatric problems in children has been mixed. Investigating this issue has been difficult due to the lack of representative longitudinal data on child mTBI that includes adequate measures of subsequent mental health symptoms and service use in young people.

**Methods:** We used data from the ABCD longitudinal cohort study to examine the association between mTBI and psychiatric diagnoses, symptoms, and psychiatric service use in over 11,000 children. In both children reporting i) previous mTBI at baseline; and ii) previously uninjured children reporting new cases of mTBI since baseline, we examined psychiatric outcomes and service use at two-year follow-up. We also compared mTBI cases to a comparison group of participants with orthopaedic injury but without mTBI. Mixed-effects models were used and adjusted for demographic and social covariates, with missing data imputed using random forest multiple imputation. To account for baseline mental health, we used propensity-score matching to identify a comparison sample matched on confounding variables and baseline outcome measures.

**Results:** When examined without adjustment for baseline mental health, both lifetime mTBI at baseline, and new occurrence of mTBI at two-year follow-up, were reliably associated with an increased risk of DSM-5 anxiety and behavioural disorders, a range of psychiatric symptom scores, and increased service use. Controlling for baseline mental health in the mTBI group using propensity-score matching eliminated all statistically reliable associations apart from anxiety disorder diagnosis and symptoms which remain associated at two-year follow-up. Evidence for association with subsequent mental health service use was inconsistent.

**Conclusion:** Anxiety was the only mental health outcome reliably associated with mTBI when baseline mental health was accounted for. Regardless of potential causality, children with mTBI are likely to present with high levels of mental health difficulties, and this remains an important comorbidity that clinicians should be aware of.

**Key points:**

- Evidence that mild TBI (mTBI) causes psychiatric problems in children has been mixed
- Longitudinal studies including measures of paediatric mTBI and pre- and post-injury psychiatric problems are required
- This longitudinal study examined the association between mTBI and psychiatric diagnoses, symptoms, and psychiatric service use in over 11,000 children aged 9-10 at baseline, and new cases of mTBI in the two years following baseline
- Higher levels of psychiatric problems and service use were associated with lifetime and new occurrence of mTBI, suggesting that paediatric mTBI should be considered a clinical marker for greater mental health attention and monitoring
- These associations were largely eliminated, or appeared equally in the orthopaedic injury comparison group, once pre-existing mental health problems were accounted for, apart from associations with anxiety and psychiatric medication use, which remained reliable. This suggests that, overall, there is limited evidence for a causal relationship between mTBI and new psychiatric problems, but that there may be evidence for a causal relationship between mTBI, anxiety problems and psychiatric medication use.

## Introduction

Traumatic brain injury (TBI) is a leading cause of death and disability worldwide (Maas et al., 2022). TBI is typically categorised into mild, moderate and severe injuries, with mild TBI (mTBI) accounting for 70-90% of all cases (Kennedy et al., 2017; Sariaslan et al., 2016). mTBI is most commonly defined as an injury resulting in a loss of consciousness for less than 30 minutes and/or posttraumatic amnesia for less than a day, and a Glasgow Coma Scale score of 13-15 (Maas et al., 2022). Incidence is particularly high in young people – in the USA, it is estimated that 1-2 million children and adolescents have a mild TBI annually (Maas et al., 2022).

Despite the majority of cases being classified as mild, the focus of research and clinical attention has traditionally been on neuropsychiatric outcomes following moderate and severe brain injury in children (Schachar et al., 2015; Stéfan & Mathé, 2016). It is well-evidenced that moderate-severe TBI in children is a risk factor for the development of psychiatric symptoms and disorders, that may persist for years following the injury (Sariaslan et al., 2016; Schachar et al., 2015). More recently, there have been concerns that important neuropsychiatric effects of mild TBI (mTBI) are being missed by both researchers and clinicians, due to many milder child TBIs not resulting in contact with the healthcare system (Ritchie & Slomine, 2022).

However, evidence for paediatric mTBI reliably raising the risk of subsequent neuropsychiatric outcomes has been mixed. A meta-analysis by Gornall et al. (2021) reported that children with concussion subsequently experienced increased internalising symptoms, externalising symptoms, and overall mental health difficulties when compared with controls. A recent retrospective cohort study of 152,321 children aged 5-18 in Canada conducted by Ledoux et al. (2022) reported concussion was associated with a subsequent increased risk of developing any mental health issue, self-harm, and psychiatric hospitalisation, compared to those who had sustained an orthopaedic injury.

Conversely, other studies have reported more equivocal results. A systematic review by the International Collaboration on Mild Traumatic Brain Injury Prognosis found no clear difference in post-concussion symptoms between children with concussion and orthopaedic controls (Keightley et al., 2014). A study of 2,160 high-school athletes by Hammer et al. (2021) found depression scores increased slightly at the 7 day assessment point but had normalised from the next assessment point (3 months) to the end of the study (12 months). Sheth et al. (2023) reported a statistically significant but weak association between mTBI and changes in sleep and behaviour in 9-10-year-olds studied as part of the Adolescent Brain Cognitive Development (ABCD) cohort study, albeit using cross-sectional data from the baseline sample.

One difficulty in interpreting these studies is that few adequately control for baseline levels of mental health outcomes that may account for reverse or shared causality for post-TBI psychiatric problems. This is important given evidence that prior mental health and behavioural problems are predictors of subsequent TBI in children, when defined as ADHD (Liou et al., 2018), Tourette syndrome (Chen et al., 2019) and any ICD-9 mental disorder (Liao et al., 2012). Furthermore, given that pre-injury mental health is strongly associated with post-injury mental health, controlling for pre-existing psychiatric outcomes is important when trying to better estimate the potential causal contribution of mTBI to the development of later psychiatric problems (Gornall et al., 2021).

Another important challenge is understanding the extent to which symptom measures adequately capture a meaningful impact of mTBI on the child and family. For example, qualitative studies have indicated that symptom measures may not sufficiently encapsulate the full range of changes to affect and functioning post-TBI (Valovich McLeod et al., 2017) or may mis-measure aspects of life that children and adolescents themselves value as important (Di Battista et al., 2015). One way of examining to what extent paediatric mTBI leads to caregiver concern regarding the young person’s mental health would be to examine help-seeking. mTBI may not lead to contact with clinical services in the majority of cases because of lack of necessity, but also potentially due to a lack of knowledge and stigma (Ritchie & Slomine, 2022). However, seeking service support for child mental health is reliably predicted by psychiatric severity and family stress (Hiscock et al., 2020; Verhulst & Der Ende, 1997), indicating that a change in help-seeking is a sensitive proxy measure of, in this cohort, parental concern about behavioural changes.

Understanding these issues at scale has been complicated, however, by the lack of representative data on child TBI, pre-existing and subsequent mental health symptoms, and service use in young people. This data has recently become available, however, in the form of the ABCD longitudinal cohort study (Feldstein Ewing et al., 2018; Volkow et al., 2018). Originally of 9– 10-year-old children, it includes structured interview assessment of TBI as well as validated measures of psychiatric symptomatology, psychiatric diagnosis, and measures of mental health service use. The ABCD study involves yearly follow-ups and representative sampling, comprising over 11,000 children from the United States.

Consequently, this study aimed to examine the association between mTBI and psychiatric symptoms, diagnoses and psychiatric service use in a cohort of children aged 9-10 at baseline and subsequently at two-year follow-up, using the ABCD cohort data.

Although this study relied on observational data, we took several measures to account for potential confounding effects. Alongside examining cross-sectional associations at baseline, we also tested whether new mTBI in children without prior mTBI was associated with changes in mental health during the 24-month follow-up period. We controlled for a range of evidence-based confounders and we included an orthopaedic injury comparison group of participants with a history of orthopaedic injury but without mTBI. Orthopaedic injury and TBI are associated with similar injury-related stress (e.g. pain, injury-related traumatic stress) and background social and behavioural predictors but only TBI is associated with damage to brain circuits and therefore a specific causal mechanism for neuropsychiatric outcomes (Kennedy et al., 2017; Ledoux et al., 2022).

Finally, we controlled for baseline levels of psychiatric outcomes. Adjusting for baseline level of the same variable used to measure outcome has been discouraged in general linear models due to evidence that it can introduce spurious statistical associations across a wide range of causal inference scenarios (Glymour et al., 2005; Lydersen & Skovlund, 2021). Here, we use propensity score matching to tackle this problem (Benedetto et al., 2018). Regression models typically address confounding factors by incorporating potential confounders as covariates to adjust for their effect. Propensity score matching estimates the treatment effect by deriving the relationship between confounders and the allocation of treatment, and creating a comparison sample matched on potential confounders (Austin et al., 2021), allowing a comparison between samples to account for baseline levels of outcomes without covariate adjustment.

## Methods

### Dataset

The ABCD Study is a prospective, longitudinal cohort study of 11,876 children recruited from 21 research sites across the United States. The ABCD sample was recruited through geographically, demographically and socioeconomically diverse school systems, employing epidemiological sampling procedures to ensure variation in sex, ethnicity/race, socioeconomic status and urbanicity that mirrors the US population (Garavan et al., 2018). Participants were aged 9-10 at baseline and the study plans to track their development through adolescence into young adulthood (Garavan et al., 2018; Karcher et al., 2018). Exclusion from enrolment included a history of severe traumatic brain injury (TBI with loss of consciousness [LOC] greater than 30 min). This study used data from ABCD Data Release 4.0, which includes measures collected from participants at baseline (n=11,876), 1-year (n=11,225), and 2-year (n=10,414) follow-up visits. Approval from the relevant Institutional Review Boards was secured prior to ABCD data collection at each site, with all parents providing written informed consent alongside assent from the participants (Clark et al., 2017). Approval for the current study was granted by the University College London Ethics Committee (Ref: CEHP/2023/593).

### Exposures

#### Lifetime mild traumatic brain injury (mTBI) at baseline

The Ohio State Traumatic Brain Injury Screen - Short Modified (OTBI) (Corrigan & Bogner, 2007). At baseline, caregivers were asked to indicate whether the child had ever experienced a TBI in their lifetime. TBI was classified according to the following OTBI summary indices: Improbable TBI (no TBI/TBI without LOC or memory loss); Possible mild TBI (TBI without LOC but memory loss); Mild TBI (TBI with LOC ≤ 30 min); Moderate TBI (TBI with LOC 30 min to 24 h); Severe TBI (TBI with LOC ≥ 24 h). Following Lopez et al. (2022), cases of moderate (n=9) and severe (n=3) TBI were removed to reduce the influence of small samples. TBI at each timepoint was categorised as a binary variable (i.e. yes/no), and only included ‘possible mild’ and ‘mild’ TBI cases.

#### New mTBI following baseline in children without prior history of mTBI

At each follow-up, caregivers were asked to indicate any new cases of TBI that had occurred since the previous study visit, using the Ohio State Traumatic Brain Injury Screen - Short Modified (OTBI). Children with new mTBI without a prior history of mTBI were identified for the second analysis.

#### Lifetime orthopaedic injury at baseline

Caregivers completed a questionnaire regarding their child’s medical history and health services utilisation. A positive response to whether the child had ever visited a doctor due to a broken bone/fracture at baseline was used to identify children with orthopaedic injury but without mTBI.

#### New orthopaedic injury following baseline in children without a prior history of orthopaedic injury

New orthopaedic injuries since the previous study visit were measured at each follow-up by parent interview and children with new orthopaedic injury without a history of orthopaedic injury or mTBI were identified.

### Outcomes

#### Child behaviour checklist (CBCL)

The CBCL is a widely used parent-rated questionnaire (Achenbach & Edelbrock, 1991) that has been used extensively to assess mental health problems following paediatric TBI (McCauley et al., 2012; Wade et al., 2020). It comprises 112 items that are aggregated into a total problem score and eight syndrome subscales (Anxious/Depressed, Withdrawn/Depressed, Somatic Complaints, Social Problems, Thought Problems, Attention Problems, Rule-breaking Behaviour and Aggressive Behaviour). For the purpose of this study, Total Problems scores were the primary outcome measure and were analysed at baseline and 2-year follow-up visit. The individual scales for externalising, internalising, anxiety, depression, attention deficit hyperactivity disorder (ADHD), conduct disorder and oppositional defiant disorder (ODD) scales at baseline and 2-year follow-up are reported as a secondary analysis.

#### DSM-5 anxiety and behavioural disorders

The caregiver-reported version of the Kiddie Schedule for Affective Disorder and Schizophrenia for DSM-5 (KSADS-5). The KSADS-5 is a semi-structured interview that indicates the presence of DSM-5 psychiatric diagnoses in children and adolescents (Kaufman et al., 1997). Any current diagnosis of general, social or separation anxiety, panic disorder or obsessive-compulsive disorder were used to identify participants in the any anxiety disorders group. The any behaviour disorders category was created by combining any positive responses to current conduct or oppositional defiant disorder diagnoses. Depression and ADHD disorders were not examined by this study, as the variables had previously been removed from the 4.0 dataset due to measurement inaccuracies.

#### Psychiatric service use

Psychiatric service use was determined by the KSADS-5 background items questionnaire. Caregivers were asked to report on whether the child had ever received i) any mental health or substance use service support, ii) psychotherapy, or iii) medication for their mental health in their lifetime (at baseline) and since the last study visit (at every follow-up).

### Covariates

#### Model 1

Caregiver reports of the child’s biological sex, age and race and ethnicity, and combined annual household income (measured as < $50,000, $50,000-$100,000 and >$100,000) were included in the first adjusted model as they have all been shown to be independent predictors of TBI and psychiatric problems (Li & Liu, 2013; Max et al., 2005; McKinlay et al., 2010). The 5-level race and ethnicity variable (Asian, Black, Hispanic, White, Other) was constructed based on a parent/caregiver report. The ‘Other’ category included those who selected more than one category (not including ‘Hispanic’), and those who selected ‘Other’ (but did not select ‘Hispanic’).

#### Model 2

Included covariates in Model 1 but additionally included neighbourhood safety, parental mental health, family conflict and traumatic experiences were included as confounders as these have shown to be independently associated with both TBI and mental health (Lopez et al., 2022; Max et al., 2005; McKinlay et al., 2010). Perceived neighbourhood safety was measured using the modified Safety/Crime Survey at baseline, where the child was asked whether they agreed with the statement “My neighbourhood is safe from crime”. Parental mental health score was created by summing self-reported DSM-5-Oriented scales of depressive, anxiety, attention deficit and antisocial personality problems by parents of children in the cohort. Family conflict score was created by summing three responses from the Family Environment Scale, where the child was asked at baseline whether they agreed with the statements “we fight a lot in our family”, “family members sometimes get so angry they throw things”, and “family members sometimes hit each other”. Experiences of childhood traumatic events (e.g. being in a car accident or being attacked) were assessed by the KSADS-5 assessment of traumatic events.

#### PSM Model

A matched sample was created that was balanced on all covariates in Models 1 and 2 and, in addition, to baseline levels of the outcome variable for each analysis.

### Analysis

Individuals were coded into three groups (mTBI, orthopaedic injury and no injury). We completed separate analyses for i) baseline association mental health outcomes and mTBI; and ii) association between mental health outcomes at age 11-12 and new mTBI / orthopaedic injury in the previous 24 months; each of which included a comparison analysis that examined the equivalent association using non-mTBI orthopaedic injury as an exposure. In the follow-up sample, only participants with new mTBI / orthopaedic injury who hadn’t experienced prior injury at baseline were included.

The association between mTBI, orthopaedic injury and each mental health outcome was measured using mixed-effects models to account for a nested structure and included study site as a random effect, with unadjusted and adjusted (see covariates section) odds ratios (OR) or beta coefficients and associated 95% confidence intervals (CI). For propensity-score matching at two-year follow-up, all new cases of mTBI and orthopaedic injury were matched to the no injury control group in a 2:1 ratio. This ratio was compared to and selected over 1:1 as it allows for better quality matches, leading to smaller standardised mean differences and reduced bias in the estimated treatment effect (Rassen et al., 2012). The matched samples created were balanced on all covariates in Models 1 and 2 (see covariates section) apart from parental mental health and, in addition, to baseline levels of the outcome variable for each analysis. Standardised mean differences of covariates before and after matching were analysed and no outliers were eliminated. Estimates were then generated from the matched cohorts using adjusted mixed-effect models (see covariates section) with OR/beta coefficients and associated 95% CI. To analyse whether propensity scores differed between injury groups, new cases of mTBI were also matched to the orthopaedic injury comparison group in a 2:1 ratio and estimates were generated using adjusted mixed-effect models. All analysis was conducted using *R* version 4.2.1 (R Core Team, 2020). Adjusted results are reported following multiple imputation. The *missForest* package in R was used to conduct random forest multiple imputation for missing variables. This method was used as it allows for simultaneous imputation of categorical and numerical variables and does not rely on distributional assumptions (Kokla et al., 2019; Shah et al., 2014). This allows for complex interactions and non-linear relations between variables, and has been shown to outperform several other imputation methods (Stekhoven & Bühlmann, 2012). The level of missing data for psychiatric outcomes at baseline and two-year follow-up prior to imputation was visualised and can be viewed in Figures S1 to S4. The percentage of missing data at baseline and two-year follow-up was 0.7% and 12.8%, respectively. The *MatchIt* package in R was used to conduct propensity score analysis from the imputed data (Ho et al., 2011). The full code for the analysis is available in full in an online archive: https://github.com/GraceRevill/pTBI-neuropsychiatric-outcomes

## Results

Baseline demographics and descriptive statistics for the sample are shown in Table 1. At baseline, 450 (3.8%) children were reported to have ever experienced mTBI, whilst 1,604 (13.5%) children had experienced orthopaedic injury in their lifetime and 9,808 (82.5%) had experienced neither. In the 2 years following baseline there were 217 (1.8%) new cases of mTBI and 466 (3.9%) new cases of orthopaedic injury in children who had not experienced prior injury. The demographics and descriptive statistics for the sample at two-year follow-up did not differ significantly from baseline and are reported in Table S1.

**Table 1.**
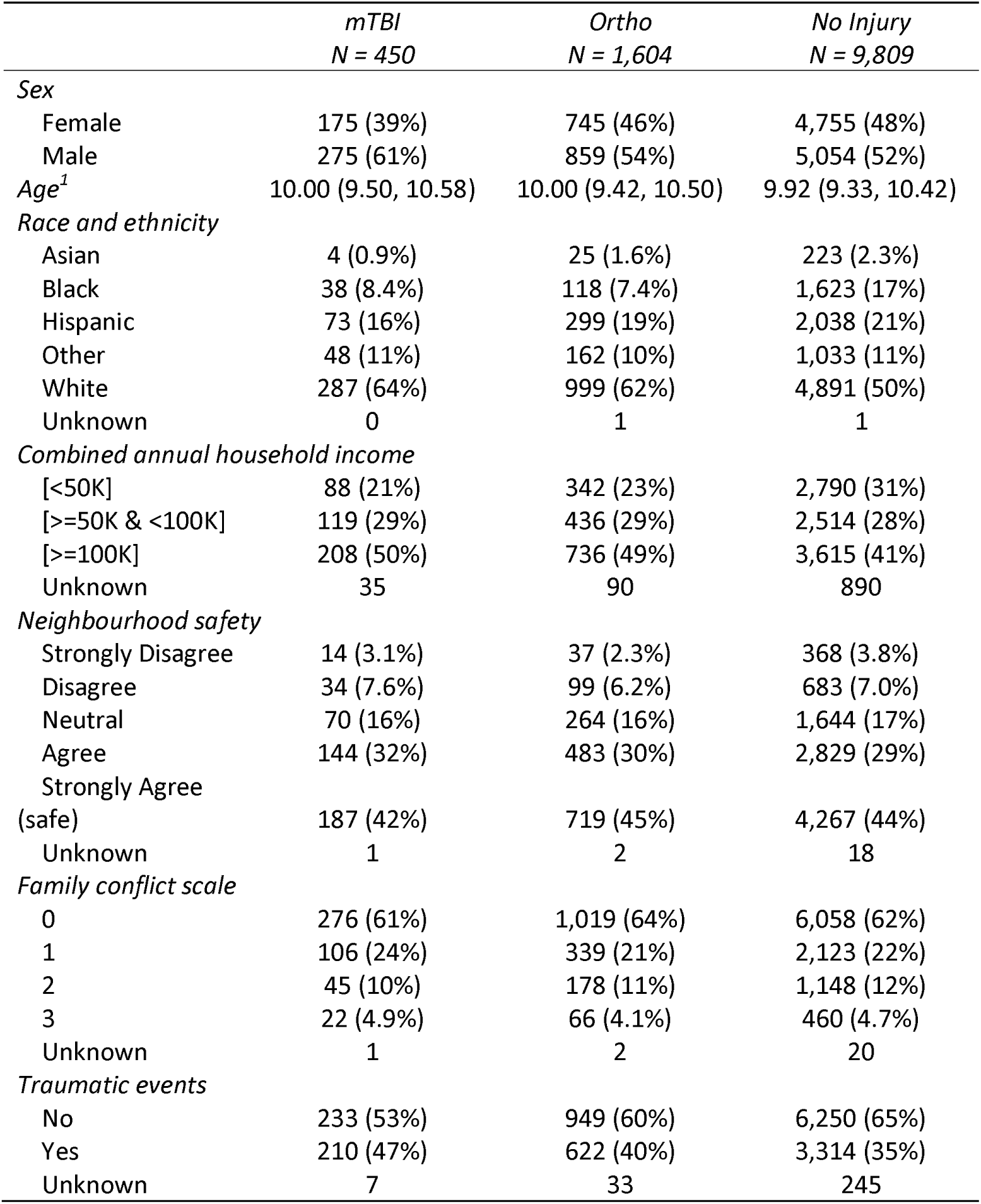
Demographics and descriptive statistics for the sample. ^1^n (%); Median (IQR)

|  | *mTBI*<br>*N = 450* | *Ortho*<br>*N = 1,604* | *No Injury*<br>*N = 9,809* |
| --- | --- | --- | --- |
| *Sex* |  |  |  |
| Female | 175 (39%) | 745 (46%) | 4,755 (48%) |
| Male | 275 (61%) | 859 (54%) | 5,054 (52%) |
| *Age*<sup>1</sup> | 10.00 (9.50, 10.58) | 10.00 (9.42, 10.50) | 9.92 (9.33, 10.42) |
| *Race and ethnicity* |  |  |  |
| Asian | 4 (0.9%) | 25 (1.6%) | 223 (2.3%) |
| Black | 38 (8.4%) | 118 (7.4%) | 1,623 (17%) |
| Hispanic | 73 (16%) | 299 (19%) | 2,038 (21%) |
| Other | 48 (11%) | 162 (10%) | 1,033 (11%) |
| White | 287 (64%) | 999 (62%) | 4,891 (50%) |
| Unknown | 0 | 1 | 1 |
| *Combined annual household income* |  |  |  |
| [<50K] | 88 (21%) | 342 (23%) | 2,790 (31%) |
| [>=50K & <100K] | 119 (29%) | 436 (29%) | 2,514 (28%) |
| [>=100K] | 208 (50%) | 736 (49%) | 3,615 (41%) |
| Unknown | 35 | 90 | 890 |
| *Neighbourhood safety* |  |  |  |
| Strongly Disagree | 14 (3.1%) | 37 (2.3%) | 368 (3.8%) |
| Disagree | 34 (7.6%) | 99 (6.2%) | 683 (7.0%) |
| Neutral | 70 (16%) | 264 (16%) | 1,644 (17%) |
| Agree | 144 (32%) | 483 (30%) | 2,829 (29%) |
| Strongly Agree |  |  |  |
| (safe) | 187 (42%) | 719 (45%) | 4,267 (44%) |
| Unknown | 1 | 2 | 18 |
| *Family conflict scale* |  |  |  |
| 0 | 276 (61%) | 1,019 (64%) | 6,058 (62%) |
| 1 | 106 (24%) | 339 (21%) | 2,123 (22%) |
| 2 | 45 (10%) | 178 (11%) | 1,148 (12%) |
| 3 | 22 (4.9%) | 66 (4.1%) | 460 (4.7%) |
| Unknown | 1 | 2 | 20 |
| *Traumatic events* |  |  |  |
| No | 233 (53%) | 949 (60%) | 6,250 (65%) |
| Yes | 210 (47%) | 622 (40%) | 3,314 (35%) |
| Unknown | 7 | 33 | 245 |

### Baseline association between mTBI and psychiatric outcomes

At baseline, 16% of children who had a history of mTBI had received an anxiety disorder diagnosis compared to 8.7% in the orthopaedic injury and 8.9% in the no injury groups. 13% of children who had a history of mTBI had a behavioural disorder diagnosis, compared to 6.8% in the orthopaedic injury and 7.1% in the no injury groups.

As can be seen from Table 2, at baseline, lifetime mTBI was reliably associated with an increased odds of psychiatric diagnosis (KSADS-5 Any anxiety disorder and KSADS-5 Any behavioural disorder) and CBCL psychiatric symptoms, which was not the case for orthopaedic injury. This association remained, although attenuated after adjustment for potential confounders. History of mTBI was positively associated with all CBCL subscales, which was not the case for orthopaedic injury, as reported in Table S2.

**Table 2.** Baseline association between mental health at age 9-10 and lifetime mTBI / orthopaedic injury. TBI = traumatic brain injury. Ortho = non-TBI orthopaedic injury.

|  | *Unadjusted*<br>*OR (95% CI)* | *Model 1*<br>*OR (95% CI)* | *Model 2*<br>*OR (95% CI)* |
| --- | --- | --- | --- |
| *KSADS-5 Any anxiety disorder* |  |  |  |
| mTBI | 1.93 (1.48 – 2.52) | 2.02 (1.54 – 2.64) | 1.61 (1.21 – 2.13) |
| Ortho injury comparison | 0.99 (0.82 – 1.20) | 1.05 (0.87 – 1.27) | 0.97 (0.79 – 1.17) |
| *KSADS-5 Any behavioural disorder* |  |  |  |
| mTBI | 2.05 (1.55 – 2.72) | 2.00 (1.50 – 2.66) | 1.56 (1.16 – 2.11) |
| Ortho injury comparison | 0.94 (0.76 – 1.15) | 0.95 (0.77 – 1.18) | 0.86 (0.69 – 1.07) |
|  | *Unadjusted*<br>*Beta (95% CI)* | *Model 1*<br>*Beta (95% CI)* | *Model 2*<br>*Beta (95% CI)* |
| *CBCL Total problems* |  |  |  |
| mTBI | 8.4 (6.70 – 10.00) | 8.39 (6.75 – 10.03) | 5.2 (3.80 – 6.60) |
| Ortho injury comparison | 0.17 (-0.77 – 1.10) | 0.44 (-0.48 – 1.36) | -0.52 (-1.30 – 0.26) |

Unadjusted and adjusted associations between baseline history of mTBI/OI and lifetime use of any mental health/substance abuse services, psychotherapy or medication are reported in Table 3. At baseline, the odds of accessing outpatient services in children who had reported mTBI were higher than in those without any injury (unadjusted OR, 2.39, 95% CI 1.86–3.07; adjusted model 1 OR, 2.39, 95% CI 1.85–3.09; adjusted model 2 OR, 1.97, 95% CI 1.51–2.56). The odds of accessing inpatient services in children who had reported mTBI were not higher than in those without any injury (unadjusted OR, 1.71, 95% CI 0.63–4.63; adjusted model 1 OR, 1.92, 95% CI 0.69–5.34; adjusted model 2 OR, 1.22, 95% CI 0.42–3.59). The proportion and percentage of psychiatric service use across injury groups at baseline is reported in Table S3.

**Table 3.** Baseline association between mental health service use at age 9-10 and lifetime mTBI / orthopaedic injury. mTBI = mild traumatic brain injury. Ortho = non-TBI orthopaedic injury. OR = odds ratio.

|  | *Unadjusted*<br>*OR (95% CI)* | *Model 1*<br>*OR (95% CI)* | *Model 2*<br>*OR (95% CI)* |
| --- | --- | --- | --- |
| *Psychotherapy* |  |  |  |
| mTBI | 1.86 (1.31 – 2.66) | 1.68 (1.18 – 2.40) | 1.47 (1.02 – 2.11) |
| Ortho injury comparison | 1.04 (0.81 – 1.34) | 0.96 (0.74 – 1.23) | 0.92 (0.71 – 1.18) |
| *Medication for mental health* |  |  |  |
| mTBI | 2.10 (1.47 – 3.00) | 1.91 (1.33 – 2.74) | 1.57 (1.08 – 2.27) |
| Ortho injury comparison | 0.78 (0.58 – 1.05) | 0.74 (0.55 – 1.01) | 0.69 (0.51 – 0.94) |
| *Any psychotherapy and/or medication* |  |  |  |
| mTBI | 1.93 (1.45 – 2.57) | 1.75 (1.31 – 2.33) | 1.48 (1.10 – 1.99) |
| Ortho injury comparison | 0.88 (0.71 – 1.09) | 0.82 (0.66 – 1.02) | 0.77 (0.62 – 0.96) |
| *Any mental health service use* |  |  |  |
| mTBI | 2.06 (1.66 – 2.55) | 1.97 (1.58 – 2.45) | 1.66 (1.33 – 2.08) |
| Ortho injury comparison | 1.00 (0.86 – 1.16) | 0.97 (0.84 – 1.13) | 0.90 (0.78 – 1.05) |

### Association between new mTBI and psychiatric outcomes at follow-up

At two-year follow-up, 13% of children who had experienced a new mTBI in the 24 months following baseline had an anxiety disorder diagnosis, compared to 7.8% in the orthopaedic injury and 7.5% in the no injury groups. 9.7% of children who had experienced mTBI in the 24 months following baseline had a behavioural disorder diagnosis at the two-year follow-up, compared to 6.7% in the orthopaedic injury and 5.1% in the no injury groups.

For mental health disorder and symptom measures, as can be seen in Table 4, without adjustment for baseline levels of outcome, new TBI was reliably associated with both KSADS-5 disorder diagnosis measures and CBCL total problems at two-year follow-up, but orthopaedic injury was not, and the association with mTBI remained after adjustment for potential confounders. New mTBI was positively associated with all CBCL subscales apart from the ODD scale, unadjusted and Model 1 depression symptoms, Model 1 ADHD symptoms, and unadjusted conduct problems, whereas orthopaedic injury was only associated with unadjusted and adjusted Model 1 internalising symptoms scores (as reported in Table S4).

**Table 4.** Association between new mTBI / orthopaedic injury and mental health at two-year follow-up. mTBI = mild traumatic brain injury. Ortho = non-TBI orthopaedic injury. PSM = propensity score matched analysis.

|  | *Unadjusted*<br>*OR (95% CI)* | *Model 1*<br>*OR (95% CI)* | *Model 2*<br>*OR (95% CI)* | *PSM Model*<br>*OR (95% CI)* |
| --- | --- | --- | --- | --- |
| *KSADS-5 Any anxiety disorder* |  |  |  |  |
| mTBI | 1.91 (1.27 – 2.88) | 2.00 (1.32 – 3.02) | 2.08 (1.36 – 3.16) | 2.23 (1.26 – 3.94) |
| Ortho injury comparison | 1.03 (0.72 – 1.48) | 1.05 (0.73 – 1.50) | 1.02 (0.71 – 1.47) | 1.12 (0.72 – 1.72) |
| mTBI vs ortho injury |  |  |  | 1.79 (1.03 – 3.06) |
| *KSADS-5 Any behavioural disorder* |  |  |  |  |
| mTBI | 2.09 (1.31 – 3.35) | 1.97 (1.23 – 3.16) | 2.12 (1.32 – 3.42) | 1.37 (0.75 – 2.45) |
| Ortho injury comparison | 1.37 (0.93 – 2.02) | 1.31 (0.89 – 1.94) | 1.28 (0.86 – 1.90) | 1.02 (0.63 – 1.60) |
| mTBI vs ortho injury |  |  |  | 1.42 (0.67 – 2.56) |
|  | *Unadjusted*<br>*Beta (95% CI)* | *Model 1*<br>*Beta (95% CI)* | *Model 2*<br>*Beta (95% CI)* | *PSM Model*<br>*Beta (95% CI)* |
| *CBCL total problems* |  |  |  |  |
| mTBI | 3.10 (1.20 – 5.10) | 3.16 (1.25 – 5.06) | 3.30 (1.50 – 5.00) | 3.4 (0.99 – 5.80) |
| Ortho injury comparison | 1.00 (-0.27 – 2.40) | 1.07 (-0.24 – 2.37) | 0.80 (-0.41 – 2.00) | 0.60 (-1.0 – 2.20) |
| mTBI vs ortho injury |  |  |  | 2.14 (-0.07 – 4.90) |

When baseline levels of mental health outcomes were accounted for using propensity score matching (Table 4), new mTBI was associated with KSADS-5 anxiety disorders at two-year follow-up, but not with behavioural disorders or CBCL total problems. Of the CBCL subscales, new mTBI was associated with CBCL anxiety at two-year follow-up (beta 0.46; 95% CI 0.11 – 0.80) but not internalising, externalising, depression, ADHD symptoms, conduct, or oppositional defiant disorder symptoms (full results in Supplementary Table 4). Orthopaedic injury was not associated with any of these measures. When we directly statistically compared new mTBI outcomes with orthopaedic injury outcomes, there were statistically reliable associations with new mTBI only for KSADS-5 anxiety diagnoses (OR 1.79; 95% CI 1.03 – 3.06) and CBCL anxiety symptoms (beta 0.38; 95% CI 0.02 – 0.74). No other disorder or symptom measures showed a statistically reliable association with mTBI when directly compared to orthopaedic injury.

For mental health service use, Table 5 reports the unadjusted and adjusted associations between the cases of mTBI / orthopaedic injury in the 12-24 months following baseline, and any mental health / substance abuse services, psychotherapy or medication that occurred within 12-24 months following baseline. Without accounting for baseline levels of outcome, in the 12-24 months following baseline, the odds of accessing outpatient services in children who had reported mTBI were significantly higher than in those children without any injury. There were no new cases of mTBI and inpatient service use in the 12-24 months following baseline. The proportion and percentage of psychiatric service use across injury groups in the 12-24 months following baseline is reported in Table S3.

**Table 5.** Association between new mTBI / orthopaedic injury and mental health service use at follow-up. mTBI = mild traumatic brain injury. Ortho = non-TBI orthopaedic injury. OR = odds ratio. PSM = Propensity Score Matched.

|  | *Unadjusted*<br>*OR (95% CI)* | *Model 1*<br>*OR (95% CI)* | *Model 2*<br>*OR (95% CI)* | *PSM Model*<br>*OR (95% CI)* |
| --- | --- | --- | --- | --- |
| *Psychotherapy* |  |  |  |  |
| mTBI | 2.23 (1.36 – 3.65) | 1.99 (1.21 – 3.26) | 2.08 (1.26 – 3.42) | 2.93 (1.42 – 6.22) |
| Ortho injury comparison | 1.50 (1.01 – 2.24) | 1.38 (0.92 – 2.07) | 1.36 (0.91 – 2.04) | 1.53 (0.91 – 2.54) |
| mTBI vs ortho injury |  |  |  | 1.42 (0.75– 2.63) |
| *Medication for mental health* |  |  |  |  |
| mTBI | 1.89 (1.14 – 3.13) | 1.74 (1.05 – 2.89) | 1.83 (1.09 – 3.05) | 1.33 (0.70 – 2.50) |
| Ortho injury comparison | 0.99 (0.62 – 1.56) | 0.93 (0.59 – 1.48) | 0.89 (0.56 – 1.42) | 1.30 (0.72 – 2.30) |
| mTBI vs ortho injury |  |  |  | 2.08 (1.03 – 4.19) |
| *Any psychotherapy and/or meds* |  |  |  |  |
| mTBI | 2.10 (1.40 – 3.15) | 1.93 (1.28 – 2.90) | 2.02 (1.34 – 3.05) | 1.80 (1.05 – 3.06) |
| Ortho injury comparison | 1.36 (0.98 – 1.89) | 1.28 (0.92 – 1.78) | 1.25 (0.89 – 1.75) | 1.63 (1.06 – 2.49) |
| mTBI vs ortho injury |  |  |  | 1.46 (0.87 – 2.34) |
| *Any mental health service use* |  |  |  |  |
| mTBI | 1.86 (1.34 – 2.57) | 1.78 (1.28 – 2.47) | 1.82 (1.30 – 2.53) | 1.64 (1.08 – 2.49) |
| Ortho injury comparison | 1.47 (1.16 – 1.87) | 1.42 (1.11 – 1.80) | 1.39 (1.09 – 1.78) | 1.34 (0.99 – 1.79) |
| mTBI vs ortho injury |  |  |  | 1.26 (0.84 – 1.87) |

When adjusted for baseline levels of outcome using propensity score matching (Table 5), new mTBI was reliably associated with ‘psychotherapy’, and associated with ‘psychotherapy and/or medication for mental health’ and ‘any mental health service use’. New orthopaedic injury was also weakly associated with psychotherapy and/or medication for mental health’. However, when compared directly with new orthopaedic injury, new mTBI was not associated with any psychiatric service use outcome other than medication for mental health, meaning no psychiatric service use outcome was reliably statistically associated with new mTBI in both comparisons against all other children or in a direct comparison to those with new orthopaedic injury.

## Discussion

Using data from a representative longitudinal cohort study of 11,876 children in the US, we report that poor mental health and higher levels of mental health service use were associated with i) lifetime mTBI at baseline, and ii) new mTBI at two years follow-up, when compared to a comparison group with orthopaedic injury. However, when baseline levels of psychiatric outcomes and service use are accounted for using a propensity score matched analysis, the statistically reliable associations largely disappear, apart from a clear associations with anxiety diagnoses and symptoms which remain reliable. There was equivocal evidence for association with mental health service use in that no psychiatric service use outcome demonstrated a statistically reliable association with new mTBI in both comparisons, namely when compared with other children and when compared to those with new orthopaedic injury. This suggests that whilst associations between mTBI and many psychiatric outcomes may be explained by pre-existing mental health problems and non-specific effect of injury, mTBI during childhood may result in an increased likelihood of experiencing anxiety.

It is unclear to what extent the observed association between mTBI and anxiety may be explained by mTBI being more anxiety-provoking than other types of injuries due to psychological factors (e.g. social challenges, concern about consequences of injury, fear of future injuries), and what may be explained by neurological damage as a direct result of the injury. It is important to note when considering the potential neurological mechanism for anxiety post-mTBI that the neurobiology of emotional regulation and perception, including anxiety, remains to be fully elucidated (Max et al., 2011). In TBI research, neuroscientific findings have suggested that TBI most likely impacts on mental health due to damage to neural networks involved in affect and behaviour. In adults, closed head injury has been shown to damage nerve fibres through shearing, leading to axonal injury and white matter damage, which can subsequently lead to cascades of neuroinflammation, vascular dysfunction, and disruption to neurotransmitter systems that are part of key circuits involved in emotion and behavioural control (Mckee & Daneshvar, 2015; Van Der Horn et al., 2020). However, clear structural abnormalities have not been consistently reported post-mTBI in children (Bigler et al., 2016; Lopez et al., 2022), meaning it is unclear to what extent similar pathophysiology is able to account for anxiety problems post paediatric mTBI, or to what extent this reflects a lack of investigation using sufficiently sensitive methodologies. Future research investigating potential mechanisms for post-mTBI anxiety, taking into consideration both psychological and neurological factors, would therefore be beneficial.

A novel aspect of this study was the inclusion of pre- and post-injury psychiatric service use in children with mTBI. Studies from different countries have shown that only a minority of children with mental health problems are in contact with psychiatric services, and that those who do receive support tend to have more severe and persistent symptoms, making it an important measure of need (Bringewatt & Gershoff, 2010; Hansen et al., 2021; Raven et al., 2017). The evidence from this study suggests that injury, rather than mTBI specifically, was a risk factor for subsequent psychiatric service use. It is important to note that this study was based in the United States which does not have a system of universal healthcare coverage and where there are large numbers of children that remain uninsured with little access to mental health services (Bornheimer et al., 2018). The extent to which these results would generalise to other countries and healthcare systems is not clear and may suggest an under-estimate of healthcare use post-injury. Future studies exploring this association in countries with different barriers to service utilisation may provide different outcomes.

This study has several important strengths, including the incorporation of multiple psychiatric domains, including psychiatric service use, in a large representative sample of children with mTBI (Garavan et al., 2018). The incorporation of a 2-year follow-up allowed for analysis of mental health outcomes in children aged 11-12 who had experienced mTBI in the previous 24 months. By including an orthopaedic injury group, where many previous studies have only used an uninjured control group, we were able to examine the extent to which associations were mTBI specific. By including propensity-score matching of new mTBI cases in the 24 months following baseline with the no injury group, we were also able to control for baseline mental health to analyse the temporality and direction of the association between mTBI and psychiatric problems. Finally, by using a demographically representative cohort sample, we expect that these findings will be more likely to generalise to the general population in the United States.

However, it is important to note some limitations of this study. The comparison with orthopaedic injury, control for a range of evidence-based confounders including pre-injury mental health using propensity-score matching and examination of association with new onset mTBI during follow-up should have reduced the chances of a confounded association, but it remains possible the results were affected by unmeasured confounding. It is important to highlight that propensity score analysis resulted in large confidence intervals which may be a reflection of the relatively small sample of mTBI cases and/or increased variability in the observed outcomes. This was not able to be tested in the current study, although the sample was matched in a 2:1, rather than a 1:1, ratio to improve estimate reliability and reduce variability. Analysis of future study visits would enable a larger mTBI sample size to investigate this issue further. It is also important to note that lifetime mTBI and psychiatric service use were retrospective measures, which may have resulted in recall bias. It was not possible to analyse the type of psychiatric medication children with mTBI were being prescribed, therefore we were not able to discern whether medication use was in direct relation to increased anxiety or another mental health problem. Furthermore, whilst the OTBI screen is a well-established, validated tool for use in adult populations, the parent proxy version has not yet been well validated in children. Additionally, it was not possible to discern the exact timepoint that lifetime or new mTBI occurred. Given that many studies have reported short-term associations of mTBI with mental health symptoms, in the weeks to months following injury, it is a possibility that mTBI did lead to psychiatric problems other than anxiety, even after controlling for baseline mental health, but that these associations had diminished by two-year follow-up. The outcomes associated with lifetime mTBI in this study may also have been affected by age at injury (Li & Liu, 2013). For example, there is some evidence to suggest younger age at injury is more predictive of anxiety symptoms (Max et al., 2011; Vasa et al., 2002) whereas older age is predictive of depressive symptoms (Max et al., 2012). Age at injury may also affect potential cognitive mediators that are associated with emotional and behavioural regulation (Li & Liu, 2013).There is some evidence to suggest that multiple TBIs may worsen outcomes including neuropsychiatric symptoms, but this full lifetime TBI history is not available with the ABCD dataset (Lopez et al., 2022; Maas et al., 2022). ABCD is a longitudinal study that will last a minimum of 10 years post-baseline, therefore analysis of future study visits will allow for a longer follow-up period and analysis of mental health trajectories in the years following injury.

Overall, this study suggests that the association between childhood mTBI and subsequent psychiatric problems and service use may be largely explained by pre-existing mental health problems and non-specific effects of injury, but that there could be evidence for a specific causal relationship between mTBI, anxiety problems and increased psychiatric medication use. It is therefore recommended that future research investigating potential mechanisms for post-mTBI anxiety, and propensity score analyses using a larger sample size and longer follow-up period are performed to elucidate this further. Regardless of causality, it remains the case that children with mTBI are likely to present with higher levels of many mental health difficulties, and this remains an important comorbidity that clinicians should be aware of. Future research should aim to elucidate the relative contribution of mTBI (e.g. damage to brain circuits), non-specific effects of injury (e.g. pain, stress) and pre-existing mental health problems in these children.

## Data Availability

Data used in the preparation of this article were obtained from the Adolescent Brain Cognitive DevelopmentSM (ABCD) Study (https://abcdstudy.org), held in the NIMH Data Archive (NDA). The full code for the analysis is available in full in an online archive: https://github.com/GraceRevill/pTBI-neuropsychiatric-outcomes

## Acknowledgements

Data used in the preparation of this article were obtained from the Adolescent Brain Cognitive Development^SM^ (ABCD) Study (https://abcdstudy.org), held in the NIMH Data Archive (NDA). This is a multisite, longitudinal study designed to recruit more than 10,000 children age 9-10 and follow them over 10 years into early adulthood. The ABCD Study® is supported by the National Institutes of Health and additional federal partners under award numbers U01DA041048, U01DA050989, U01DA051016, U01DA041022, U01DA051018, U01DA051037, U01DA050987, U01DA041174, U01DA041106, U01DA041117, U01DA041028, U01DA041134, U01DA050988, U01DA051039, U01DA041156, U01DA041025, U01DA041120, U01DA051038, U01DA041148, U01DA041093, U01DA041089, U24DA041123, U24DA041147. A full list of supporters is available at https://abcdstudy.org/federal-partners.html. A listing of participating sites and a complete listing of the study investigators can be found at https://abcdstudy.org/consortium_members/. ABCD consortium investigators designed and implemented the study and/or provided data but did not necessarily participate in the analysis or writing of this report. This manuscript reflects the views of the authors and may not reflect the opinions or views of the NIH or ABCD consortium investigators. The ABCD data repository grows and changes over time. The ABCD data used in this report came from https://doi.org/10.15154/mpdm-9w22

## Abbreviations

CBCL: child behaviour checklist
KSADS-5: Kiddie Schedule for Affective Disorder and Schizophrenia for DSM-5
mTBI: mild traumatic brain injury.

