## Supplementary material for "Only Anxiety Remains Reliably Associated with Paediatric Mild Traumatic Brain Injury at Two Years Follow-up After Adjusting for Pre-Existing Mental Health"

|  | *mTBI*  *N = 217* | *Ortho*  *N = 466* | *No Injury*  *N = 10,533* | |
| --- | --- | --- | --- | --- |
| *Sex* |  |  |  | |
| Female | 87 (40%) | 221 (47%) | 5,040 (48%) | |
| Male | 130 (60%) | 245 (53%) | 5,492 (52%) | |
| Unknown | 0 | 0 | 1 | |
| *Age* | 12.08 (11.50, 12.50) | 12.00 (11.42, 12.58) | 12.00 (11.42, 12.58) | |
| Unknown | 4 | 28 | 1,007 | |
| *Race and Ethnicity* |  |  |  | |
| Asian | 3 (1.4%) | 7 (1.5%) | 231 (2.2%) | |
| Black | 17 (7.8%) | 32 (6.9%) | 1,547 (15%) | |
| Hispanic | 30 (14%) | 80 (17%) | 2,111 (20%) | |
| Other | 29 (13%) | 45 (9.7%) | 1,098 (10%) | |
| White | 138 (64%) | 302 (65%) | 5,544 (53%) | |
| Unknown | 0 | 0 | 2 | |
| *Combined annual household income* | | | | |
| [<50K] | 35 (17%) | 85 (20%) | | 2,812 (29%) |
| [>=50K & <100K] | 67 (32%) | 129 (30%) | | 2,741 (28%) |
| [>=100K] | 105 (51%) | 216 (50%) | | 4,110 (43%) |
| Unknown | 10 | 36 | | 870 |
| *Neighbourhood safety* | | | |  |
| Strongly Disagree | 5 (2.3%) | 9 (1.9%) | | 370 (3.5%) |
| Disagree | 13 (6.0%) | 23 (4.9%) | | 703 (6.7%) |
| Neutral | 32 (15%) | 67 (14%) | | 1,770 (17%) |
| Agree | 74 (34%) | 139 (30%) | | 3,060 (29%) |
| Strongly Agree (safe) | 93 (43%) | 228 (49%) | | 4,614 (44%) |
| Unknown | 0 | 0 | | 16 |
| *Family conflict scale* | | | |  |
| 0 | 146 (67%) | 283 (61%) | | 6,554 (62%) |
| 1 | 40 (18%) | 109 (23%) | | 2,274 (22%) |
| 2 | 23 (11%) | 53 (11%) | | 1,209 (11%) |
| 3 | 8 (3.7%) | 20 (4.3%) | | 480 (4.6%) |
| Unknown | 0 | 1 | | 16 |
| *Traumatic events* | |  | |  |
| No | 129 (61%) | 281 (62%) | | 6,618 (64%) |
| Yes | 84 (39%) | 171 (38%) | | 3,667 (36%) |
| Unknown | 4 | 14 | | 248 |

**Supplementary Table 1.** Demographics and descriptive statistics for the two-year follow-up sample

^1^n (%); Median (IQR)

|  | *Unadjusted*  *Beta (95% CI)* | *Model 1*  *Beta (95% CI)* | *Model 2*  *Beta (95% CI)* |
| --- | --- | --- | --- |
| *Internalising symptoms score* |  |  |  |
| mTBI | 2.20 (1.70 – 2.70) | 2.23 (1.72 – 2.74) | 1.30 (0.87 – 1.80) |
| Ortho injury comparison | 0.17 (-0.12 – 0.46) | 0.16 (-0.13 – 0.45) | -0.12 (-0.37 – 0.13) |
| *Externalising symptoms score* |  |  |  |
| mTBI | 2.20 (1.70 – 2.80) | 2.26 (1.72 – 2.80) | 1.40 (0.93 – 1.90) |
| Ortho injury comparison | 0.00 (-0.31 – 0.30) | 0.13 (-0.17 – 0.44) | -0.12 (-0.39 – 0.15) |
| *Anxiety symptoms score* |  |  |  |
| mTBI | 0.74 (0.51 – 0.97) | 0.75 (0.52 – 0.97) | 0.39 (0.19 – 0.59) |
| Ortho injury comparison | 0.06 (-0.06 – 0.19) | 0.06 (-0.07 – 0.18) | -0.05 (-0.17 – 0.06) |
| *Depression symptoms score* |  |  |  |
| mTBI | 0.77 (0.58 – 0.96) | 0.76 (0.57 – 0.95) | 0.47 (0.30 – 0.64) |
| Ortho injury comparison | 0.01 (-0.10 – 0.11) | 0.01 (-0.09 – 0.12) | -0.07 (-0.17 – 0.02) |
| *ADHD symptoms score* |  |  |  |
| mTBI | 1.20 (0.88 – 1.40) | 1.10 (0.86 – 1.40) | 0.73 (0.48 – 0.98) |
| Ortho injury comparison | -0.01 (-0.17 – 0.14) | 0.04 (-0.12 – 0.19) | -0.08 (-0.22 – 0.06) |
| *Conduct problems score* |  |  |  |
| mTBI | 0.73 (0.51 – 0.95) | 0.77 (0.55 – 0.98) | 0.49 (0.29 – 0.70) |
| Ortho injury comparison | -0.04 (-0.16 – 0.08) | 0.04 (-0.08 – 0.16) | -0.04 (-0.15 – 0.07) |
| *ODD symptoms score* |  |  |  |
| mTBI | 0.70 (0.51 – 0.89) | 0.67 (0.49 – 0.86) | 0.40 (0.23 – 0.58) |
| Ortho injury comparison | 0.02 (-0.08 – 0.13) | 0.03 (-0.08 – 0.14) | -0.05 (-0.15 – 0.05) |

**Supplementary Table 2.** Association between mental health symptoms at age 9-10 and lifetime mTBI / orthopaedic injury. TBI = traumatic brain injury. Ortho = non-TBI orthopaedic injury.

| *Baseline* | *mTBI*  *N = 450* | *Ortho*  *N = 1,604* | *No Injury*  *N = 9,808* |
| --- | --- | --- | --- |
| Psychotherapy | 36 (8.0%) | 75 (4.7%) | 440 (4.5%) |
| Medication for mental health | 36 (8.0%) | 50 (3.1%) | 391 (4.0%) |
| Outpatient support | 83 (18%) | 139 (8.7%) | 814 (8.3%) |
| Inpatient support | 4 (0.9%) | 5 (0.3%) | 51 (0.5%) |
| Any mental health service use | 124 (28%) | 246 (15%) | 1,505 (15%) |
| *Two-year follow-up* | *mTBI*  *N = 217* | *Ortho*  *N = 466* | *No Injury*  *N = 10,533* |
| Psychotherapy | 18 (8.5%) | 27 (6.2%) | 410 (4.3%) |
| Medication for mental health | 17 (8.0%) | 20 (4.6%) | 455 (4.8%) |
| Outpatient support | 27 (13%) | 44 (10%) | 789 (8.3%) |
| Inpatient support | 0 (0.0%) | 1 (0.2%) | 50 (0.5%) |
| Any mental health service use | 48 (23%) | 85 (19%) | 1,383 (15%) |

**Supplementary Table 3.** Proportion and percentage of psychiatric service use across injury groups at baseline and two-year follow-up.

|  | *Unadjusted*  *Beta (95% CI)* | *Model 1*  *Beta (95% CI)* | *Model 2*  *Beta (95% CI)* | *PSM Model*  *Beta (95% CI)* |
| --- | --- | --- | --- | --- |
| *Internalising symptoms score* |  |  |  |  |
| mTBI | 1.00 (0.40 – 1.70) | 1.03 (0.40 – 1.66) | 1.10 (0.46 – 1.60) | 0.74 (-0.12 – 1.6) |
| Ortho injury comparison | 0.48 (0.05 – 0.92) | 0.44 (0.00 – 0.87) | 0.37 (-0.04 – 0.77) | 0.43 (-0.09 – 0.95) |
| mTBI vs ortho injury |  |  |  | 0.52 (-0.35 – 1.40) |
| *Externalising symptoms score* |  |  |  |  |
| mTBI | 0.75 (0.13 – 1.4) | 0.80 (0.18 – 1.47) | 0.84 (0.25 – 1.40) | 0.64 (-0.15 – 1.4) |
| Ortho injury comparison | 0.27 (-0.15 – 0.70) | 0.33 (-0.10 – 0.75) | 0.24 (-0.16 – 0.65) | 0.00 (-0.54 – 0.53) |
| mTBI vs ortho injury |  |  |  | 0.65 (-0.14 – 1.4) |
| *Anxiety symptoms score* |  |  |  |  |
| mTBI | 0.53 (0.26 – 0.79) | 0.53 (0.27 – 0.79) | 0.53 (0.28 – 0.79) | 0.46 (0.11 – 0.80) |
| Ortho injury comparison | 0.15 (-0.03 – 0.33) | 0.12 (-0.06 – 0.30) | 0.10 (-0.07 – 0.27) | 0.05 (-0.17 – 0.27) |
| mTBI vs ortho injury |  |  |  | 0.38 (0.02 – 0.74) |
| *Depression symptoms score* |  |  |  |  |
| mTBI | 0.25 (-0.01 – 0.50) | 0.24 (-0.02 – 0.49) | 0.25 (0.01 – 0.49) | 0.29 (-0.03 – 0.62) |
| Ortho injury comparison | 0.13 (-0.04 – 0.30) | 0.11 (-0.06 – 0.29) | 0.09 (-0.08 – 0.25) | 0.04 (-0.19 – 0.26) |
| mTBI vs ortho injury |  |  |  | 0.12 (-0.21 – 0.45) |
| *ADHD symptoms score* |  |  |  |  |
| mTBI | 0.34 (0.02 – 0.65) | 0.30 (-0.01 – 0.61) | 0.32 (0.02 – 0.61) | 0.40 (-0.01 – 0.80) |
| Ortho injury comparison | -0.01 (-0.23 – 0.20) | 0.00 (-0.21 – 0.21) | -0.04 (-0.24 – 0.17) | -0.10 (-0.37 – 0.17) |
| mTBI vs ortho injury |  |  |  | 0.38 (-0.04 – 0.80) |
| *Conduct problems score* |  |  |  |  |
| mTBI | 0.23 (-0.02 – 0.48) | 0.26 (0.02 – 0.51) | 0.27 (0.03 – 0.51) | 0.24 (-0.09 – 0.58) |
| Ortho injury comparison | 0.12 (-0.06 – 0.29) | 0.15 (-0.02 – 0.32) | 0.13 (-0.04 – 0.29) | 0.15 (-0.05 – 0.35) |
| mTBI vs ortho injury |  |  |  | 0.19 (-0.13 – 0.51) |
| *ODD symptoms score* |  |  |  |  |
| mTBI | 0.16 (-0.06 – 0.39) | 0.16 (-0.07 – 0.38) | 0.17 (-0.04 – 0.38) | 0.18 (-0.10 – 0.46) |
| Ortho injury comparison | 0.06 (-0.10 – 0.21) | 0.07 (-0.09 – 0.21) | 0.03 (-0.11 – 0.18) | -0.04 (-0.23 – 0.15) |
| mTBI vs ortho injury |  |  |  | 0.12 (-0.18 – 0.42) |

**Supplementary Table 4.** Association between mental health symptoms at age 11-12 and new mTBI / orthopaedic injury in the previous 24 months. TBI = traumatic brain injury. Ortho = non-TBI orthopaedic injury. PSM = propensity score matched.

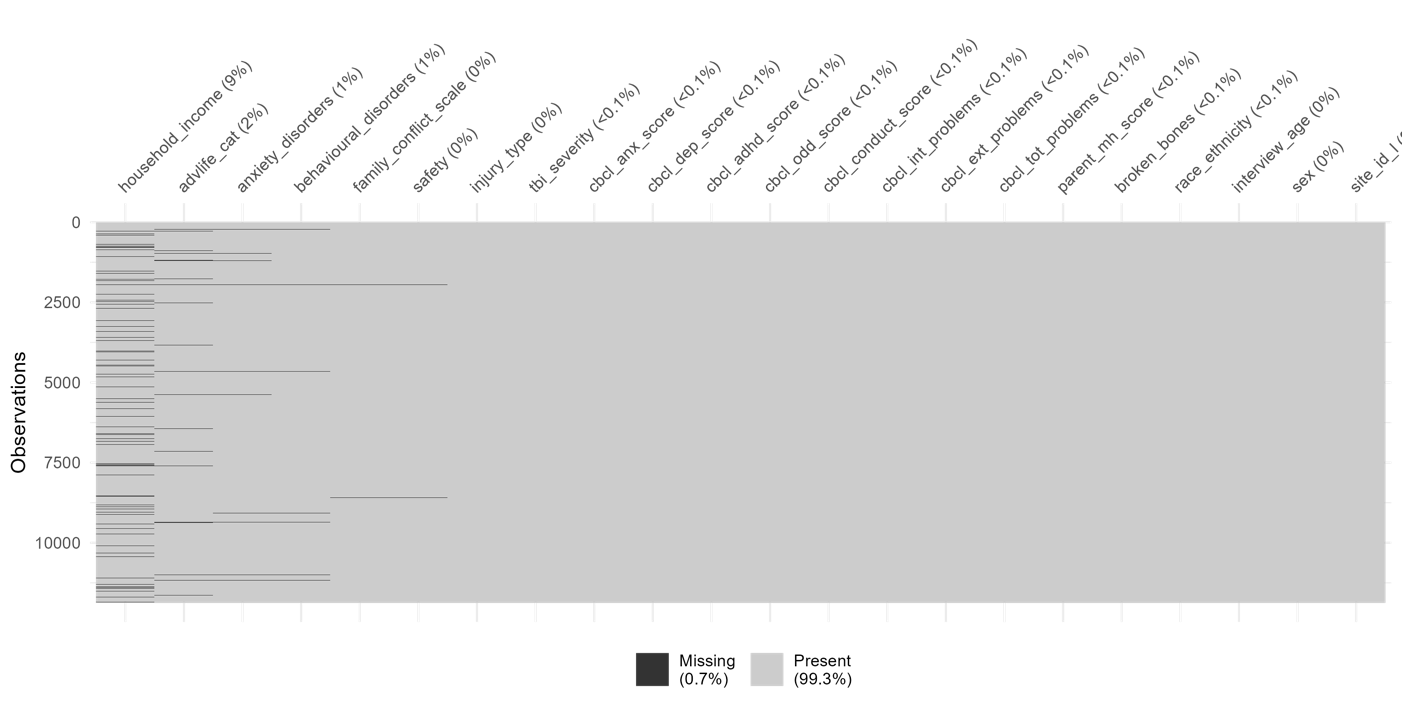

**Supplementary Figure 1.** Visualisation of missing data for psychiatric disorders and symptoms at baseline

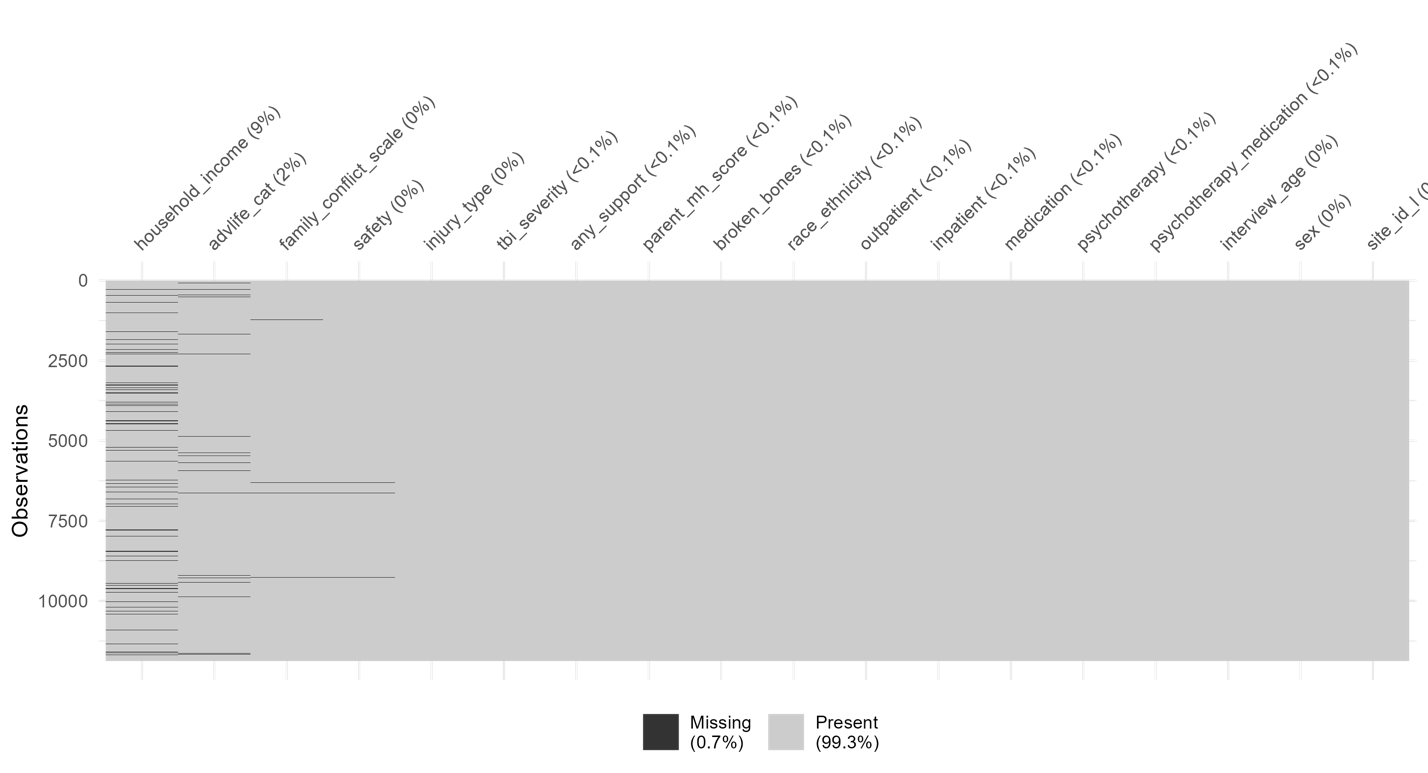

**Supplementary Figure 2.** Visualisation of missing data for psychiatric service use at baseline

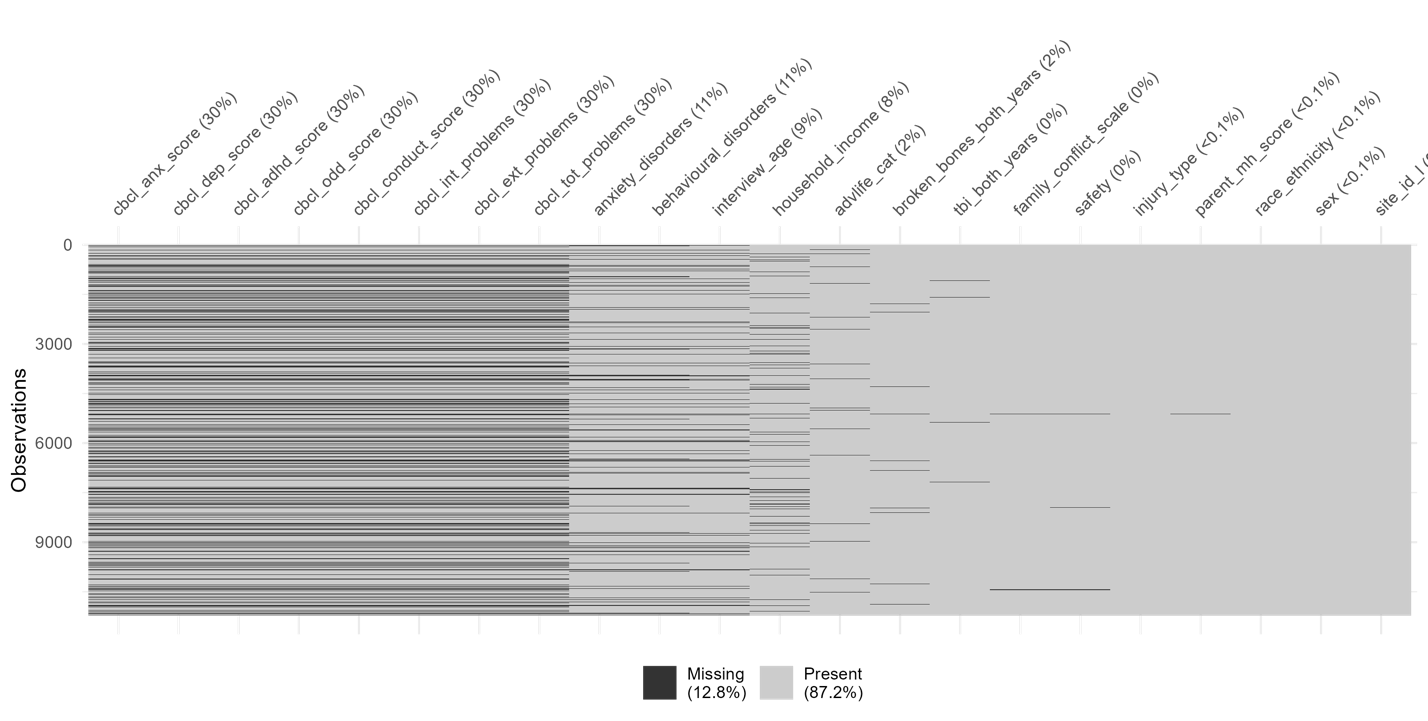

**Supplementary Figure 3.** Visualisation of missing data for psychiatric disorders and symptoms at two-year follow-up

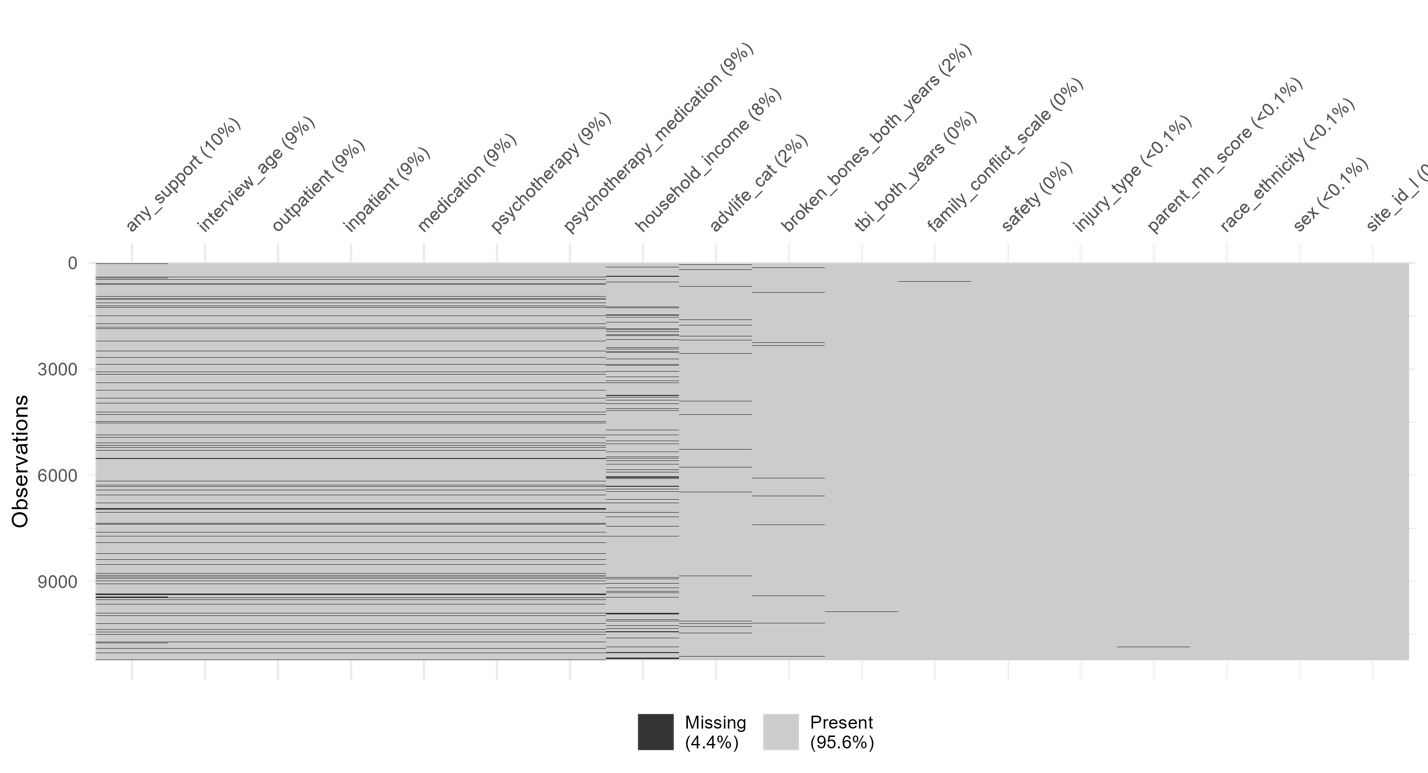

**Supplementary Figure 4.** Visualisation of missing data for psychiatric service use at two-year follow-up
